# Excess Mortality and Years of Potential Life Lost Among Black People in the US from 1999 to 2020

**DOI:** 10.1101/2022.11.12.22282253

**Authors:** César Caraballo, Daisy S. Massey, Chima D. Ndumele, Trent Haywood, Shayaan Kaleem, Terris King, Yuntian Liu, Yuan Lu, Marcella Nunez-Smith, Herman A. Taylor, Karol E. Watson, Jeph Herrin, Clyde W. Yancy, Jeremy S. Faust, Harlan M. Krumholz

## Abstract

**Importance:** Amid efforts in the United States to promote health equity, there is a need to assess progress in reducing excess deaths and years of potential life lost (YPLL) among Black people compared with White people.

**Objective:** To evaluate trends in excess mortality and YPLL among Black people compared with White people.

**Design:** Serial cross-sectional.

**Setting:** National data from the Centers for Disease Control and Prevention, 1999-2020

**Participants:** Non-Hispanic White and non-Hispanic Black people

**Exposures:** Race as documented in the death certificates.

**Main outcomes and measures:** Excess age-adjusted all-cause and disease-specific mortality rate (per 100,000 individuals) and YPLL among Black people compared with White people.

**Results:** From 1999 to 2020, the total number of excess deaths was 658,356 and 1,154,108 among Black females and males, representing 34,938,070 and 47,005,048 excess YPLL among Black females and males. The excess deaths and YPLL were largest among infants and non-elderly adults. Heart disease had the most excess deaths. From 1999, the age-adjusted excess mortality rate declined at an annual average of -9.0 (95% CI: -10.0, -8.0; P<0.001) until 2015 among Black women and at an annual average of -16.3 (95% CI: -20.9, -11.6; P<0.001) until 2012 among Black men, followed by no significant change until 2019 in either group. From 2019 to 2020, excess deaths increased from 90.4 to 192 per 100,000 Black women and from 209.8 to 395 per 100,000 Black men, reaching rates approximating those of 1999. The trends in rates of excess YPLL followed a similar pattern.

**Conclusions and relevance:** Over a recent 22-year period, Black people in the US lost more than 80 million years of life when compared with White people. After a period of progress, improvements stalled, and most gains were eliminated in 2020.

**KEY POINTS:** *Question:* How many excess deaths and years of potential life lost (YPLL) for Black people, compared with White people, occurred in the United States from 1999 through 2020?

*Findings:* Based on Centers for Disease Control and Prevention data, excess deaths and YPLL persisted throughout the period, with initial progress followed by little improvement, and then worsening in 2020 to about 1999 levels. Black people had 1.8 million excess deaths and over 80 million YPLL over the study period.

*Meaning:* After initial progress, excess mortality and YPLL among Black people stagnated and then worsened, indicating a need for new approaches.

## INTRODUCTION

In 1985, the U.S. Department of Health and Human Services (HHS) Secretary Margaret M. Heckler issued the Report of the Secretary’s Task Force on Black and Minority Health, also known as the Heckler Report.^1^ The report found that Black people had strikingly higher mortality rates than White people, resulting in almost 60,000 excess deaths a year relative to White People. Race offers no intrinsic biological reason for those categorized as Black individuals to have worse outcomes than White individuals. The most likely reasons for the differences are the burden of acquired risk factors, influence of social determinants of health, limitations in access to care and structural barriers indicative of bias (i.e., structural racism).^2-9^

Though these mortality disparities are well documented, what remains absent is a unifying metric that can be used for accountability and improvement. The Heckler report focused on excess mortality, as had a prior seminal academic report.^10^ However, the United States government did not adopt excess death as an official metric of health equity for accountability. Comparative death rates are a common way to describe health inequity but are not a summary measure and do not convey the impact of differences in mortality at different age groups. Quantifying excess years of potential life lost (YPLL) provides a deeper measure of the differential burden of excess mortality as a function of race by giving more weight to deaths at younger ages, thus addressing the impact on the social and economic loss from early deaths.^11,12^ This metric calls attention to the aggregate burden of health inequities, and addressing this knowledge gap potentially motivates national reporting of excess deaths and YPLL among Black people. Moreover, communicating the mortality inequity within a population in terms of excess number of people lost, or the excess years lost from mortality creates the more compelling case for change. However, there is little recent information on the trends in YPLL by race, and especially so in Black populations compared with White populations.

Accordingly, we propose excess deaths and YPLL as candidate metrics to track and drive change, as they convey the effect of health inequities on lives and years lost, placing in bright relief the health consequences of race in the United States. We calculated, over the most recent 22-year period for which data are available, the difference in age-adjusted excess mortality and YPLL for Black people compared with White people in the U.S. We also investigated these disparities by causes of death, and how they varied by age. We focus on the period before the COVID-19 pandemic, but also report results for 2020, which provide an early perspective on differences attributable to the pandemic. These metrics can draw attention on the magnitude of the disparities, attracting resources, and promoting action to address the unmet need for progress.^13^

## METHODS

### Data source

We used national U.S. death certificates data from the Centers for Disease Control and Prevention Wide-ranging Online Data for Epidemiologic Research (CDC WONDER) for years 1999 to 2020.^14^ For this period, we obtained the annual life expectancy by 5-year age groups from the National Center for Health Statistics (NCHS) life tables.^15^ The Yale Institutional Review Boards waived this study from review.

### Study population

We included data from non-Hispanic Black (Black) and non-Hispanic White (White) people aged <85 years old. From CDC WONDER, we obtained annual number of deaths, population size (N), and age-adjusted mortality rate (per 100,000 individuals) by race (Black, White), sex (female, male), age (years), and cause of death (eMethods and eTable 1).

Race and ethnicity were determined by the information recorded in the death certificates, which is reported by the funeral director as provided by an informant (e.g., next of kin) or on the basis of observation if there is no informant available.^14^ Though fraught with the potential for significant inaccuracies, prior work demonstrates near complete agreement for race ascertainment among non-Hispanic White or non-Hispanic Black.^16,17^

### Statistical analysis

For all analyses we stratified by sex, and rates were reported per 100,000 individuals. For each year, we first calculated excess mortality rate by subtracting the year-specific age-adjusted mortality rate among White people from that of Black people. Then, we estimated annual YPLL (defined as the number of years that a person would have lived should they not died when they did) rate for each race by multiplying each 5-year age group crude mortality rate by their respective annual sex- and age-specific life expectancy in years and obtaining the mean rate across all age groups.^11^ For both racial groups, we used year-, age-, and sex-specific White life expectancy as the referent metric (eTable 2).^15^ We then estimated the Black-White age-adjusted mortality rate ratio, YPLL rate ratio, and the cumulative 22-year total number of excess deaths and excess YPLL among Black people (eMethods).

To assess the trends over the study period, we graphically assessed the relationship between each metric and study year. Based on this assessment, we used autoregressive integrated moving average models using a 1-year correlation and modelled time as a linear spline with knots that reflected the observed inflection points (eMethods). Separately, we calculated and visually evaluated annual rates in age-adjusted excess deaths and YPLL by the leading causes of death (eMethods and eTable 1).

Finally, to estimate mortality differences by age, we combined all study years and used the average crude mortality rate for each 5-year age group and subtracted the mortality rate of White people from that of Black people of the same age group. Similarly, to estimate the YPLL rate differences by age, we estimated the annual crude mortality rate for each age group, multiplied it by their respective annual sex- and age-specific life expectancy in years, and then estimated the mean YPLL rate for each age group across the entire study period.

Cause of death heatmaps were produced using Python version 3.7.^18-22^ All statistical tests performed were two-sided, with a level of significance of 0.05, using Stata SE version 17.0 (StataCorp, College Station TX).

## RESULTS

### Cumulative 22-year Excess Mortality and YPLL Among Black People

From 1999 to 2020, when compared with the mortality rate of White females and males, the mean annual excess mortality rate among Black females and males was 148.6 and 284.2 deaths per 100,000 individuals, respectively, representing an estimated total of 658,356 excess deaths among Black females and 1,154,108 excess deaths among Black males over the study period (eFigure 1). The mean 22-year excess YPLL rate per 100,000 individuals was 7932 among females and 11579 among males, representing an estimated total of 34,938,070 excess YPLL among Black females and 47,005,048 excess YPLL among Black males (eFigure 2).

### Temporal Trends in Racial Differences in Mortality and YPLL Rates

From 1999 to 2011, the age-adjusted excess mortality rate declined from 404 to 211 excess deaths per 100,000 individuals among Black males, at an annual average estimated decline of -16.3 excess deaths per 100,000 individuals (95% CI: -20.9, -11.6; P<0.001; Figure 1).

**Figure 1.**
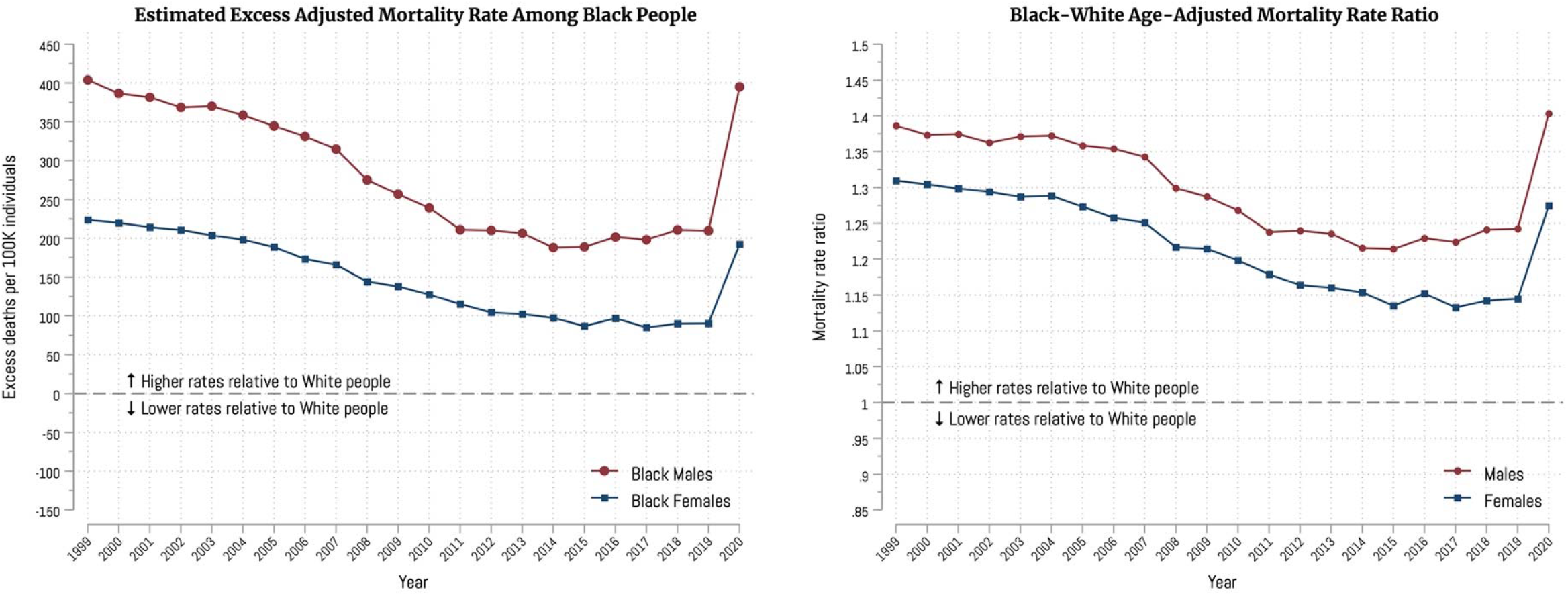
Age-Adjusted Excess Mortality Rate (Left Panel) and Rate Ratio (Right Panel) Among Black People, 1999-2020 Left panel displays the annual excess age-adjusted mortality rates among Black people, defined as the difference in each year’s mortality rate between Black people and White people. Right panel displays the annual age-adjusted mortality rate ratio, obtained by dividing the annual mortality rate among Black people by that of White people. An excess mortality rate of zero represents the absence of a difference, and a rate ratio of one represents equal rates between both groups. Rates were estimated per 100,000 individuals.

However, the rate plateaued from 2012 to 2019 (-1.4 per year; 95% CI: -9.6, +6.7; P=0.73), with a subsequent single-year increase to 395 excess deaths per 100,000 individuals in 2020– coinciding with the early pandemic experiences; reaching rates close to those of 20 years before. Among Black females, the estimated age-adjusted excess mortality rate declined from 224 excess deaths (per 100,000 individuals) in 1999 to 87 in 2015, at an annual average estimated decline of -9.0 (95% CI: -10.0, -8.0; P<0.001) excess deaths. There was no progress from 2016 to 2019 (+0.9 per year; 95% CI: -3.7, +5.5; P=0.70), and rates increased to 192 excess deaths per 100,000 individuals in 2020; similar to rates of 2005 (Figure 1). The age-adjusted mortality rate ratio between the Black and White people ranged from 1.13 to 1.31 among females and from 1.21 to 1.40 among males over the 22-year period (Figure 1).

In 1999, the excess YPLL for Black males and Black females was 14,964 and 10,806 per 100,000 people, respectively. There was a continuous decline until 2011 among males, reaching 9348 excess YPLL per 100,000 individuals, and until 2012 among females, reaching 6210 excess YPLL per 100,000 individuals (P<0.001 for males and females; Figure 2). Then there was a plateau until 2019 for both groups (P=0.90 for males; P=0.89 for females), after which the excess YPLL among both Black males and Black females increased in 2020 to rates similar to those of 19 and 13 years before, respectively (Figure 2). The annual YPLL rate ratio followed a similar pattern over the 22-year period (Figure 2).

**Figure 2.**
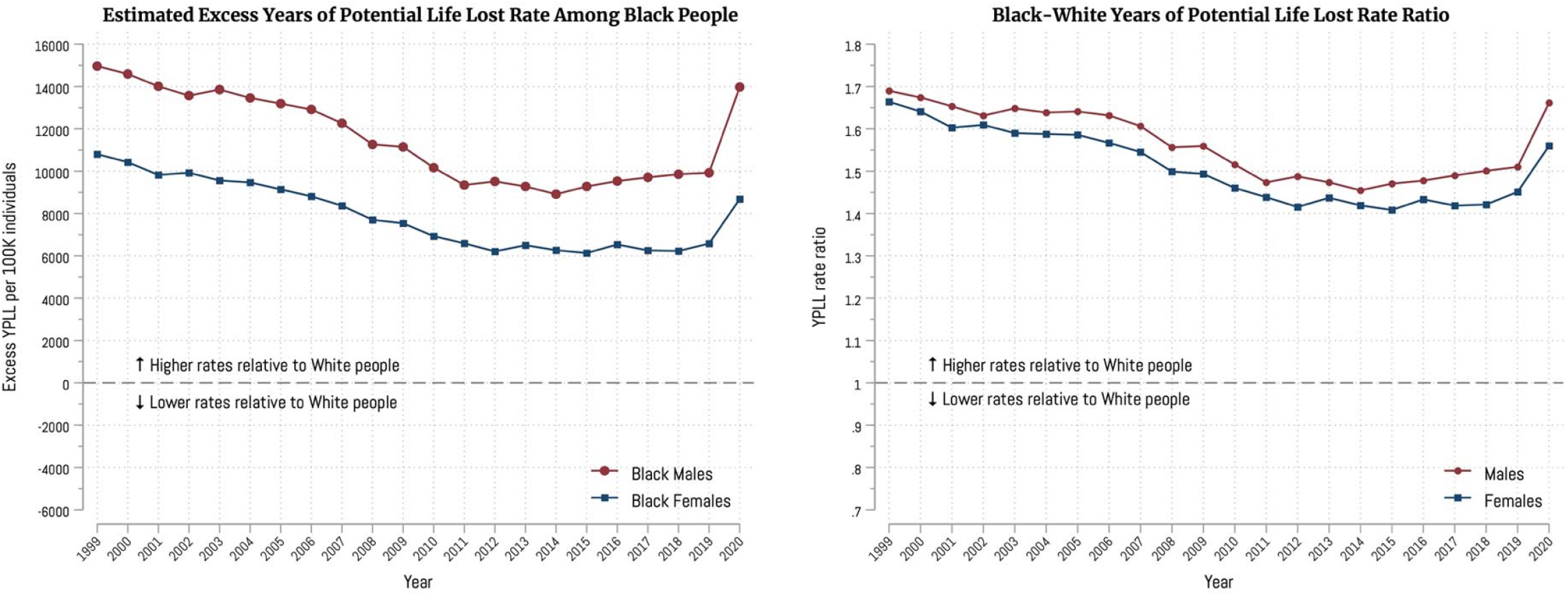
Excess Years of Potential Life Lost Rate (Left Panel) and Rate Ratio (Right Panel) Among Black People, 1999-2020 Left panel displays the annual excess YPLL rates among Black people, defined as the difference in each year’s YPLL rate between Black people and White people. Right panel displays the annual YPLL rate ratio, obtained by dividing the annual YPLL rate among Black people by that of White people. An excess YPLL rate of zero represents the absence of a difference, and a rate ratio of one represents equal rates between both groups. Rates were estimated per 100,000 individuals. Abbreviations: YPLL, years of potential life lost.

When analyzed by cause of death, the highest excess mortality rates among Black people through the study period were due to heart disease (Figure 3); in both sex groups there was a gradual decrease in mortality due to heart disease in the decade of 2010, metric that increased again in 2020. Among males, there was a gradual reduction in excess mortality rates due to cancer, diabetes, and HIV, whereas excess mortality due to assault and cerebrovascular diseases were persistently high over the study period, with an increase in 2020. Among females, the excess mortality rates due to cancer, diabetes, and cerebrovascular disease gradually decreased from 1999 to 2020. There were consistent patterns in YPLL rates by cause of death for females and males except for perinatal deaths, which had the highest YPLL rates through the study period (Figure 3).

**Figure 3.**
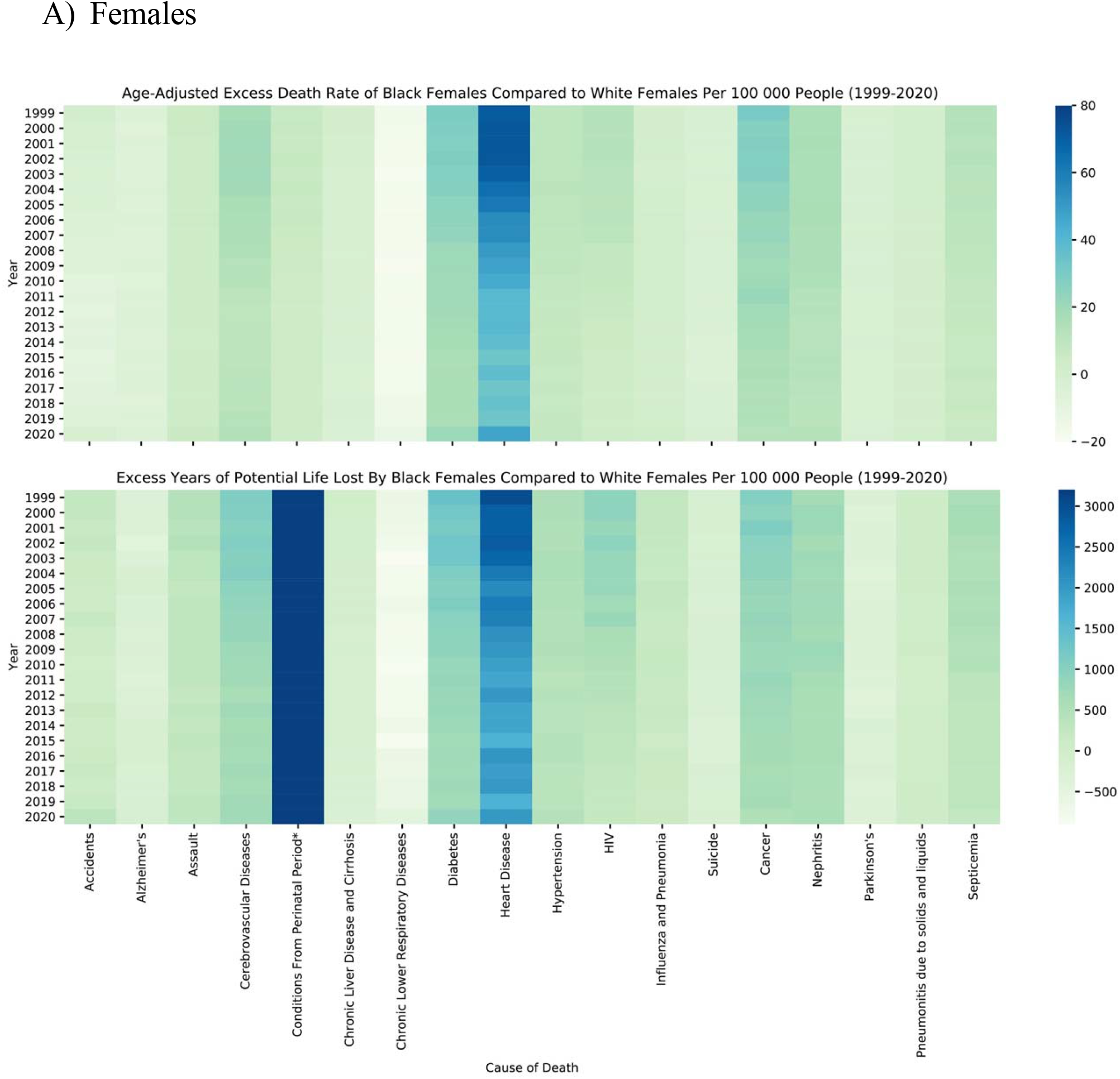
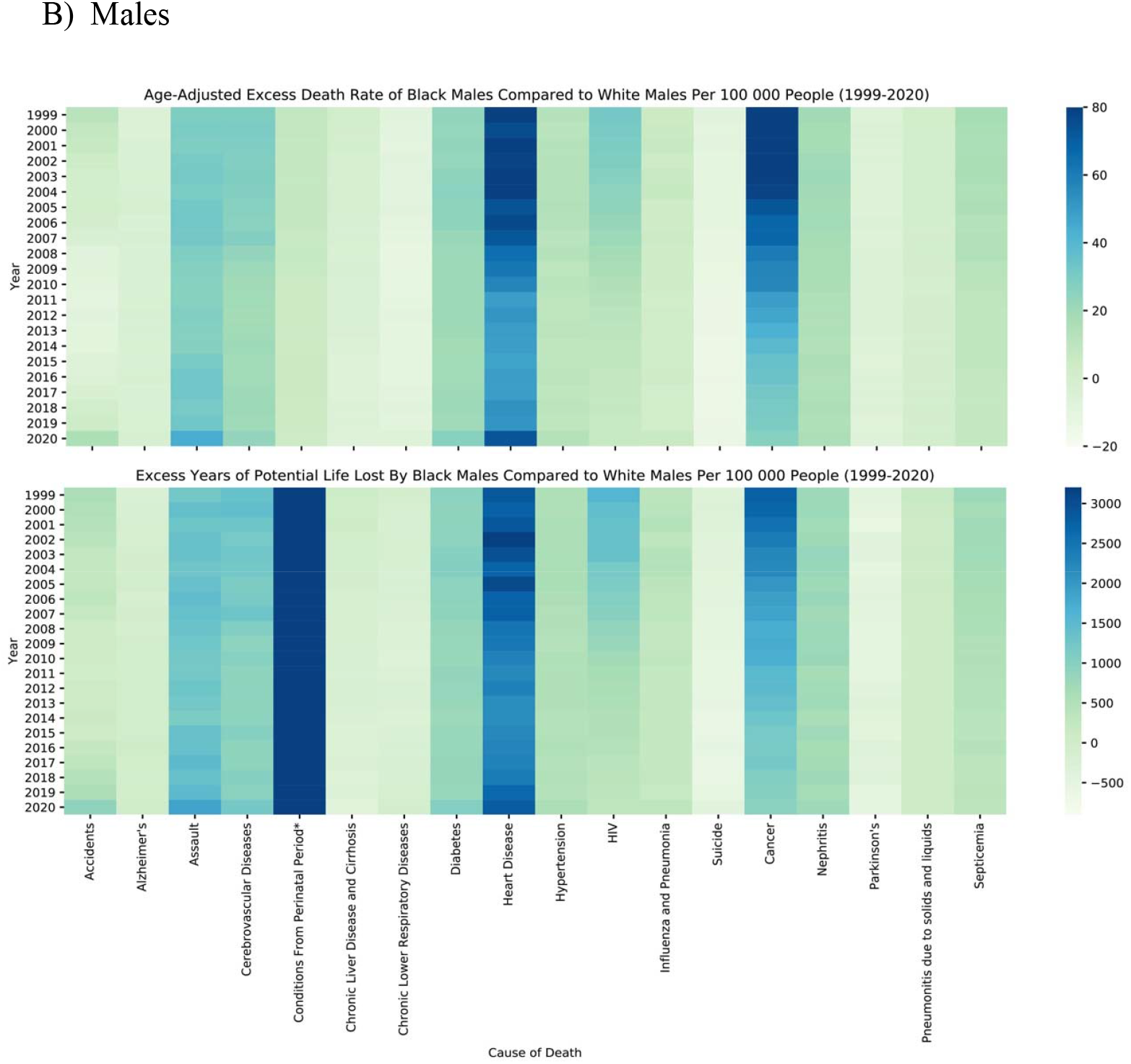
Black People Excess Mortality and Years of Potential Life Lost Rates by Cause of Death, 1999-2020 Top panels display annual excess age-adjusted mortality rates among Black females and Black males. Bottom panels display annual excess years of potential life lost among Black people. Excess mortality and YPLL rates were defined as the difference in each year’s mortality rate (or YPLL rate) between Black people and White people. A higher number in both estimates represent a socially negative outcome; zero represents the absence of a difference. Presented here are the 22-year 15 leading causes of death among White people and among Black people, separately, resulting in a total of 18 unique causes of death (eMethods). All rates are estimated per 100,000 individuals. Information related to COVID-19 deaths are not displayed in the figure because data were only applicable to a single year. Abbreviations: YPLL, years of potential life lost.

In 2020, the excess age-adjusted mortality rates due to COVID-19 were 47 per 100,000 Black females and 80 per 100,000 Black males; similarly, the excess YPLL (per 100,000 individuals) due to COVID-19 were 1759 among Black females and 2572 among Black males.

### Racial Differences in Mortality Rate and YPLL Rate by Age Group

The relationship between excess mortality rates and age among females and males is shown in Figure 4. Among Black females and Black males younger than 1 year, there were 654 and 776 excess deaths per 100,000 individuals, respectively. Such rates narrowed among those aged 1 and 14 years. Among males, the excess mortality rate increased with age starting among those 15 years and older, reaching 1302 excess deaths per 100,000 individuals among those between 75-79 years. A similar pattern, although at a smaller magnitude, was observed among Black females. The increase in mortality rate difference increased among Black females older than 20 years and reached a maximum of 677 excess deaths per 100,000 individuals among those between 75-79 years old. For both males and females, there was a slight decrease in excess deaths among those aged 80-84 years. In relative terms, patterns for the mortality rate ratio between Black and White individuals were similar (Figure 4).

**Figure 4.**
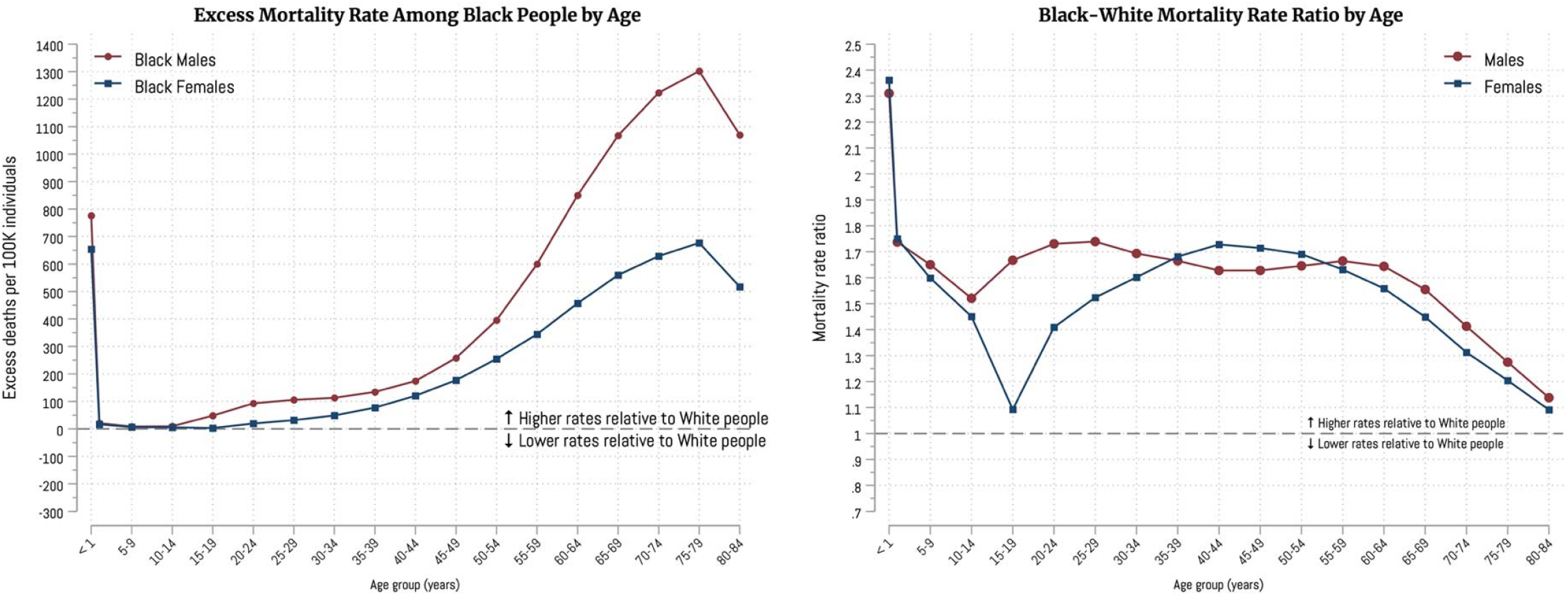
Excess Mortality Rate (Left Panel) and Rate Ratio (Right Panel) Among Black People by Age Group Left panel displays the excess mortality rates among Black people by age group, defined as the difference in each age group mortality rate between Black people and White people. Right panel displays the mortality rate ratio by age group, obtained by dividing each age group mortality rate among Black people by that of White people. An excess mortality rate of zero represents the absence of a difference, and a rate ratio of one represents equal rates between both groups. Rates were estimated per 100,000 individuals.

The greatest difference in YPLL rate between Black people and White people was among the youngest (Figure 5). Among those younger than 1 year old, excess YPLL rate was 59,232 per 100,000 Black males and 53,061 per 100,000 Black females, decreasing to its minimum among those between 1 and 14 years old. Consistent with the excess mortality pattern, the excess YPLL rate increased with age among males older than 15 years and among females older than 20 years, peaking among those aged 65-69 years old and decreasing among those older. The YPLL rate ratio across age groups followed a similar pattern (Figure 5).

**Figure 5.**
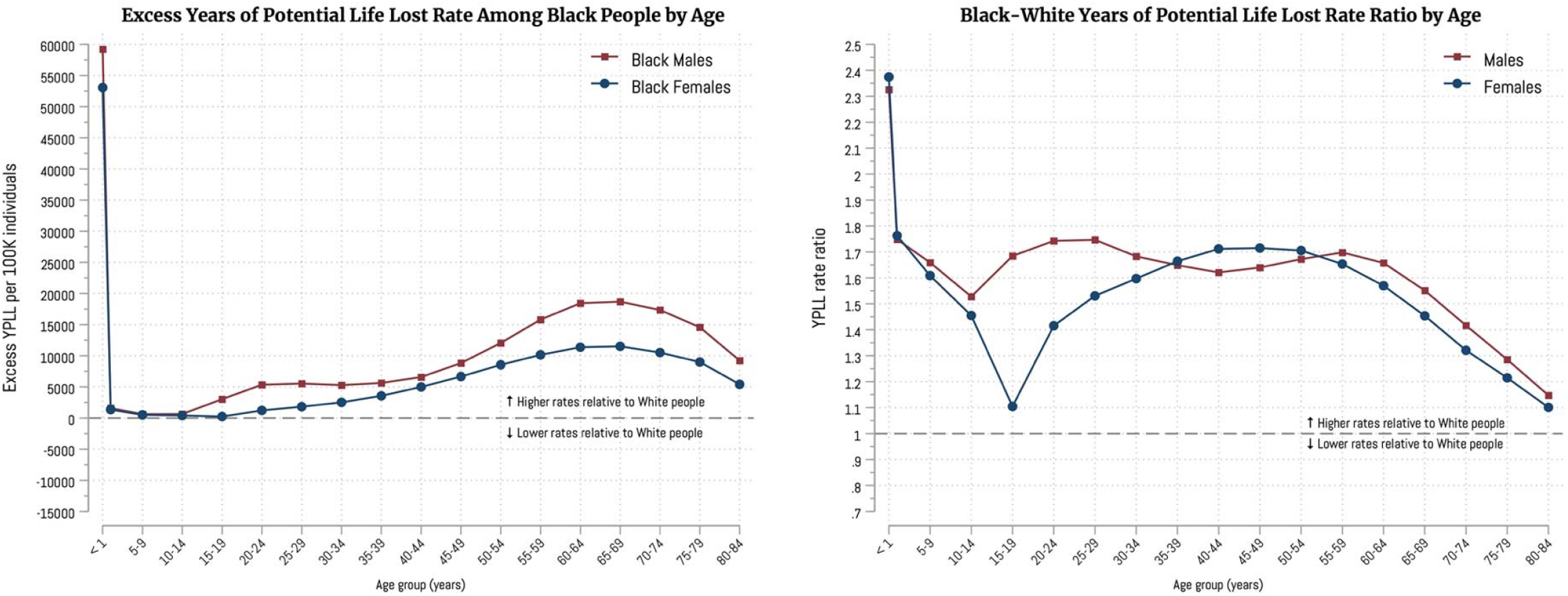
Excess Years of Potential Life Lost Rate (Left Panel) and Rate Ratio (Right Panel) Among Black People by Age Group Left panel displays the excess YPLL rates among Black people by age group, defined as the difference in each age group YPLL rate between Black people and White people. Right panel displays the YPLL rate ratio by age group, obtained by dividing each age group YPLL rate among Black people by that of White people. An excess YPLL rate of zero represents the absence of a difference, and a rate ratio of one represents equal rates between both groups. Rates were estimated per 100,000 individuals. Abbreviations: YPLL, years of potential life lost.

## DISCUSSION

In this study of US mortality data from 1999 to 2020, we estimate that there were 35 million excess YPLL among Black females and 47 million excess YPLL among Black males. After a period of progress, improvements stalled and even reversed abruptly coincident with the first year of the pandemic, when the number of excess deaths was about the same as 1999, and the YPLL loss was even greater. Moreover, even at the nadir of excess of deaths and YPLL in 2012, there were more than 2 million excess years of potential life lost among Black people compared with White people. This loss of life was most prominent among infants, with Black-White mortality and YPLL rate ratios greater than 2.3 among those <1 year old. Heart disease in females and males and cancer in males had the most excess deaths. The implication is that current efforts to curb or eliminate mortality disparities are failing, and progress, when made, is fragile.

This study quantifies the numbers of years lost to Black populations in the United States. This substantial difference in deaths and YPLL requires further study and importantly requires more precision as to cause of death and concomitant risk factors. However, the contribution of unmeasured social factors largely related skin color, a characteristic with little intrinsic biological meaning but immense social consequences, is the major presumed but unsubstantiated cause. Accounting for certain specific risks attributable to genetic ancestry, the absence of biological determinants, the role of social variables–including, social determinants of health, access to care, structural barriers, and subconscious bias–are leading candidate causal factors.^2-5^ Of note, this study shows a brief period of progress that stalled and then was erased at the onset and well into the early pandemic. The findings demonstrate the potential for progress but indicate the fragility of the gains and herald a need for new approaches. As such, an annual publicly reported metric of race-based YPLL may be useful for national accountability.

Prior studies have found persistently large numbers of excess deaths among Black people but have not explicitly quantified excess deaths or YPLL, nor established a usable metric for year over year comparison. For example, a study found that the mortality rate disparity between Black people and White people remained mostly stable from 1960 to 2000,^23^ and another report estimated that 2,687,051 excess deaths occurred among Black people from 1970 to 2004.^24^ We found that excess mortality declined substantially from 1999 through the early 2010s, consistent with other studies of Black and White mortality rates.^25,26^ This observation could indicate the success of efforts towards reducing disparities. However, it may also be paradoxically explained by disproportionally higher increases in mortality among White adults in the late 2000s.^27,28^ Importantly, the health disparity in life expectancy, which had been narrowing in the 2000s, stagnated and may have begun to widen in 2015.^29,30^ Our finding reveal that the disparity in mortality rates between Black and White people did not improve further up to 2019.

Another key observation is the marked increase in excess Black mortality during the first year of the COVID-19 pandemic. This is a novel finding because this study places it in context of excess mortality and YPLL trends–a useful metric should another pandemic-type public health calamity occur. The increase in excess mortality during the pandemic was so large that it eliminated the gains achieved in reducing mortality differences with White people during the prior two decades. Such a marked reversal underscores the potential perniciousness of social factors disproportionally positioning Black people at a higher risk of adverse health outcomes, including barriers to health care, higher multimorbidity prevalence, and worse average health status.^6,7,31^ It also indicates the recent gains did not insulate Black people from losing those gains amid the pandemic.

The excess deaths and YPLL were elevated among most of the major causes of death, though heart disease was the most prominent. This is consistent with the findings from Kyalwazi and colleagues^32^ who found that, despite a decline in the absolute difference, age-adjusted cardiovascular mortality rates were persistently higher among Black people when compared with White people from 1999 to 2019. It is likely that disparities in rates of hypertension control and other factors associated with heart disease are in part responsible for this excess loss of life.^33,34^ Cancer was also an important source of the disparity. Importantly, heart disease and cancer have many shared modifiable risk factors and are importantly affected by social determinants of health. Targeted and renewed efforts aimed at diseases that disproportionately drive differences are necessary to make sure clinical advancements are experienced uniformly across the population.

We note the sobering disparity among infants and during childhood, accounting for a markedly elevated number of excess deaths and an even more pronounced disparity in YPLL. This excess mortality occurred in a period of life of highest vulnerability and calls for new dedicated public health initiatives targeting early childhood health. After childhood, the excess deaths and YPLL among Black people became evident in early adulthood and generally increased with age. These excess deaths in working age groups may be the most socially disruptive^12^ given the destabilizing effect on productivity and economic gains for Black families, potentially contributing to the perpetuation of generational racial disparities. Among those older than age 65 years these disparity metrics declined, as would be expected because of the persistently lower average lifespan among Black people.^30,35^

The implications of these findings are important. Black people, regardless of cause or the burden of risk factors, in the US continue to die at much greater rates than White people, with a measurable excess loss of years of life. These metrics, especially YPLL, are suitable for public reporting and may inspire strategies directed toward more in-depth root cause analysis and where qualified by evidence of efficacy, implementation of steps to improve these disparities. Excess mortality and YPLL by race could serve as a major national gauge of progress towards achieving health equity. Moreover, such measures might also have value at the State and local level.

Our study has limitations. First, there are known inaccuracies in race and ethnicity information reported in death certificates^16,36^. However, we can be confident in information about death, the focus of our study. Moreover, in prior studies there is near complete agreement between self-reported race and the race reported in non-Hispanic Black and White decedent’s death certificates.^17^ The cause of death may be uncertain in many cases, especially during the pandemic, but that does not detract from our central findings of a sustained major difference in mortality, measured in millions of lives and even more millions of years of life lost, between Black and White people in the U.S over multiple decades.

In conclusion, in the US, excess deaths and YPLL among Black people persist and by scale warrant national attention and indicate the need for new strategies. Over a recent 22-year period, Black people in America lost more than 80 million years of life. While encouraging gains were noted in the early 2000s, those gains were eliminated by 2020 and YPLL replicated 1999 benchmarks. The call to action generated nearly 40 years ago from the Heckler Report remains extant today. With millions more lives and life-years at stake, new strategies are needed.

## Supporting information

eMethods; eTable 1; eTable 2; eFigure 1; eFigure 2

## Data Availability

All data used in this study is publicly available from https://wonder.cdc.gov/. All the code files used to analyze the data are publicly available from https://zenodo.org/record/7278473 Dr. Caraballo had full access to all the data in the study and takes responsibility for the integrity of the data and the accuracy of the data analysis.

https://zenodo.org/record/7278473

## Data availability statement

All data used in this study are publicly available from https://wonder.cdc.gov/. All the code files used to analyze the data are publicly available from https://zenodo.org/record/7278473 Dr. Caraballo had full access to all the data in the study and takes responsibility for the integrity of the data and the accuracy of the data analysis.

