## Supplementary material for "Excess Mortality and Years of Potential Life Lost Among Black People in the US from 1999 to 2020": eMethods; eTable 1; eTable 2; eFigure 1; eFigure 2

**TABLE OF CONTENTS**

**eMethods**

**eTable 1.** ICD-10 group codes for cause of death

**eTable 2**. White people life expectancy by age group and year

**eFigure 1**. Annual and cumulative total number of excess deaths among Black people

**eFigure 2**. Annual and cumulative total number of excess years of potential life lost among Black people

**eMethods**

*Cause of death*

In the CDC WONDER data, the underlying cause-of-death is defined as "the disease or injury which initiated the train of events leading directly to death, or the circumstances of the accident or violence which produced the fatal injury,"^1^ and is determined by the information entered by the physician on the death certificate. Causes of death are classified in accordance with the International Classification of Disease Tenth Revision (ICD-10).^1^ For the cause of death section of this study, we included the top 15 leading causes of death among White people and the 15 leading causes of death among Black people, separately, resulting in a total of 18 unique causes of death. For 2020 only, we also included deaths due to COVID-19.

*Statistical analysis*

To obtain relative metrics, we also estimated the age-adjusted Black–White mortality rate ratio by dividing the Black people age-adjusted mortality rate by the corresponding White people age-adjusted mortality rate. Using the same approach, we also estimated the Black–White YPLL rate ratio.

To estimate the cumulative 22-year total number of excess deaths and excess YPLL among Black people, we multiplied each year’s estimated excess mortality rate and excess YPLL rate–obtained as described above–by their respective annual Black population (N), and added the results over the study period.

To assess trends over time, we modelled time as a linear spline with knots that reflected the observed inflection points over years. For excess mortality rates, these inflections points were 2011 and 2019 for males, and 2015 and 2019 for females. For excess YPLL rates, the knots were 2011 and 2019 for males, and 2012 and 2019 for females. To account for the serial correlation of annual rates, we used autoregressive integrated moving average models using a 1-year correlation.

**eTable 1.** ICD-10 Codes for single underlying cause of death

| **Cause of Death** | **ICD-10 Codes** |
| --- | --- |
| Accidents | V01 - X59, Y85 - Y86 |
| Alzheimer’s | G30 |
| Assault | U01 - U02, X85 - Y09, Y87.1 |
| Cancer | C00 - C97 |
| Cerebrovascular Diseases | I60 - I69 |
| Certain conditions originating in the perinatal period | P00-P96 |
| Chronic Liver Disease and Cirrhosis | K70, K73 - K74 |
| Chronic Lower Respiratory Diseases | J40 - J47 |
| Diabetes | E10 - E14 |
| Heart Disease | I00 - I09, I11, I13, I20 - I51 |
| Hypertension | I10, I12, I15 |
| HIV | B20 - B24 |
| Influenza and Pneumonia | J09 - J18 |
| Nephritis | N00 - N07, N17 - N19, N25 - N27 |
| Parkinson’s | G20 - G21 |
| Pneumonitis due to Solids and Liquids | J69 |
| Septicemia | A40 - A41 |
| Suicide | U03, X60 - X84, Y87.0 |

**eTable 2**. White females (A) and males (B) life expectancy by age group and year

A) Females

| **Age,**  **years** | **Life expectancy (years)** | | | | | | | | | | | | | | | | | | | | | |
| --- | --- | --- | --- | --- | --- | --- | --- | --- | --- | --- | --- | --- | --- | --- | --- | --- | --- | --- | --- | --- | --- | --- |
|  | **1999** | **2000** | **2001** | **2002** | **2003** | **2004** | **2005** | **2006** | **2007** | **2008** | **2009** | **2010** | **2011** | **2012** | **2013** | **2014** | **2015** | **2016** | **2017** | **2018** | **2019** | **2020** |
| **< 1** | 79.9 | 80 | 80.2 | 80.3 | 80.4 | 80.8 | 80.4 | 80.6 | 80.6 | 80.7 | 81.1 | 81.1 | 81.1 | 81.2 | 81.2 | 81.2 | 81 | 81 | 81 | 81.1 | 81.3 | 80.2 |
| **1-4** | 79.3 | 79.4 | 79.6 | 79.7 | 79.8 | 80.2 | 79.8 | 80 | 80 | 80.2 | 80.4 | 80.5 | 80.4 | 80.6 | 80.5 | 80.6 | 80.4 | 80.4 | 80.3 | 80.4 | 80.6 | 79.5 |
| **5-9** | 75.4 | 75.5 | 75.7 | 75.8 | 75.9 | 76.3 | 75.8 | 76.1 | 76.1 | 76.2 | 76.5 | 76.6 | 76.5 | 76.6 | 76.6 | 76.6 | 76.4 | 76.5 | 76.4 | 76.5 | 76.6 | 75.6 |
| **10-14** | 70.5 | 70.5 | 70.8 | 70.8 | 71 | 71.3 | 70.9 | 71.1 | 71.1 | 71.3 | 71.5 | 71.6 | 71.5 | 71.6 | 71.6 | 71.6 | 71.5 | 71.5 | 71.4 | 71.5 | 71.7 | 70.6 |
| **15-19** | 65.5 | 65.6 | 65.8 | 65.9 | 66 | 66.4 | 65.9 | 66.1 | 66.2 | 66.3 | 66.6 | 66.6 | 66.6 | 66.7 | 66.7 | 66.7 | 66.5 | 66.5 | 66.4 | 66.6 | 66.7 | 65.6 |
| **20-24** | 60.7 | 60.7 | 60.9 | 61 | 61.1 | 61.5 | 61 | 61.3 | 61.3 | 61.4 | 61.7 | 61.7 | 61.7 | 61.8 | 61.7 | 61.8 | 61.6 | 61.6 | 61.5 | 61.6 | 61.8 | 60.7 |
| **25-29** | 55.8 | 55.8 | 56.1 | 56.1 | 56.3 | 56.6 | 56.2 | 56.4 | 56.4 | 56.5 | 56.8 | 56.9 | 56.8 | 56.9 | 56.9 | 56.9 | 56.7 | 56.8 | 56.7 | 56.8 | 56.9 | 55.9 |
| **30-34** | 50.9 | 50.9 | 51.2 | 51.2 | 51.4 | 51.8 | 51.3 | 51.5 | 51.6 | 51.7 | 52 | 52 | 52 | 52.1 | 52 | 52.1 | 51.9 | 52 | 51.9 | 52 | 52.1 | 51.1 |
| **35-39** | 46.1 | 46.1 | 46.3 | 46.4 | 46.6 | 46.9 | 46.5 | 46.7 | 46.8 | 46.9 | 47.2 | 47.2 | 47.2 | 47.3 | 47.2 | 47.3 | 47.2 | 47.2 | 47.2 | 47.3 | 47.4 | 46.4 |
| **40-44** | 41.3 | 41.3 | 41.6 | 41.6 | 41.8 | 42.1 | 41.7 | 41.9 | 42 | 42.1 | 42.4 | 42.5 | 42.4 | 42.5 | 42.5 | 42.5 | 42.4 | 42.5 | 42.5 | 42.6 | 42.7 | 41.7 |
| **45-49** | 36.6 | 36.6 | 36.9 | 36.9 | 37.1 | 37.4 | 37 | 37.2 | 37.3 | 37.4 | 37.7 | 37.8 | 37.7 | 37.8 | 37.8 | 37.9 | 37.8 | 37.9 | 37.8 | 37.9 | 38 | 37.1 |
| **50-54** | 32 | 32 | 32.3 | 32.4 | 32.5 | 32.9 | 32.4 | 32.6 | 32.7 | 32.9 | 33.2 | 33.2 | 33.2 | 33.3 | 33.3 | 33.3 | 33.2 | 33.3 | 33.3 | 33.3 | 33.5 | 32.6 |
| **55-59** | 27.5 | 27.5 | 27.8 | 27.9 | 28 | 28.4 | 27.9 | 28.2 | 28.3 | 28.4 | 28.7 | 28.8 | 28.8 | 28.8 | 28.8 | 28.9 | 28.8 | 28.9 | 28.8 | 28.9 | 29 | 28.2 |
| **60-64** | 23.2 | 23.2 | 23.5 | 23.6 | 23.7 | 24.1 | 23.6 | 23.8 | 24 | 24.1 | 24.4 | 24.4 | 24.5 | 24.5 | 24.6 | 24.7 | 24.5 | 24.7 | 24.6 | 24.7 | 24.8 | 24 |
| **65-69** | 19.2 | 19.2 | 19.5 | 19.5 | 19.7 | 20 | 19.5 | 19.8 | 19.8 | 20 | 20.3 | 20.3 | 20.3 | 20.4 | 20.4 | 20.5 | 20.4 | 20.5 | 20.5 | 20.6 | 20.7 | 20 |
| **70-74** | 15.5 | 15.5 | 15.7 | 15.8 | 15.9 | 16.2 | 15.7 | 15.9 | 16 | 16.1 | 16.4 | 16.4 | 16.4 | 16.5 | 16.5 | 16.6 | 16.5 | 16.6 | 16.6 | 16.6 | 16.7 | 16.1 |
| **75-79** | 12.1 | 12.1 | 12.3 | 12.3 | 12.5 | 12.8 | 12.1 | 12.3 | 12.4 | 12.6 | 12.9 | 12.8 | 12.8 | 12.9 | 12.9 | 13 | 12.9 | 13 | 12.9 | 13 | 13.1 | 12.5 |
| **80-84** | 9.1 | 9.1 | 9.3 | 9.3 | 9.4 | 9.7 | 9.1 | 9.3 | 9.3 | 9.4 | 9.7 | 9.6 | 9.6 | 9.6 | 9.6 | 9.7 | 9.6 | 9.8 | 9.7 | 9.7 | 9.8 | 9.3 |

B) Males

| **Age,**  **years** | **Life expectancy (years)** | | | | | | | | | | | | | | | | | | | | | |
| --- | --- | --- | --- | --- | --- | --- | --- | --- | --- | --- | --- | --- | --- | --- | --- | --- | --- | --- | --- | --- | --- | --- |
|  | **1999** | **2000** | **2001** | **2002** | **2003** | **2004** | **2005** | **2006** | **2007** | **2008** | **2009** | **2010** | **2011** | **2012** | **2013** | **2014** | **2015** | **2016** | **2017** | **2018** | **2019** | **2020** |
| **< 1** | 74.6 | 74.8 | 75 | 75.1 | 75.3 | 75.7 | 75.4 | 75.7 | 75.8 | 75.9 | 76.3 | 76.4 | 76.4 | 76.5 | 76.5 | 76.5 | 76.3 | 76.2 | 76.1 | 76.2 | 76.3 | 75 |
| **1-4** | 74.1 | 74.3 | 74.5 | 74.6 | 74.8 | 75.2 | 74.9 | 75.1 | 75.3 | 75.4 | 75.7 | 75.8 | 75.9 | 75.9 | 75.9 | 75.9 | 75.7 | 75.6 | 75.5 | 75.6 | 75.7 | 74.3 |
| **5-9** | 70.2 | 70.3 | 70.6 | 70.7 | 70.9 | 71.3 | 71 | 71.2 | 71.4 | 71.5 | 71.8 | 71.9 | 71.9 | 72 | 72 | 72 | 71.8 | 71.6 | 71.6 | 71.7 | 71.8 | 70.4 |
| **10-14** | 65.3 | 65.4 | 65.6 | 65.7 | 65.9 | 66.3 | 66 | 66.3 | 66.4 | 66.5 | 66.9 | 67 | 67 | 67.1 | 67 | 67 | 66.9 | 66.7 | 66.6 | 66.7 | 66.8 | 65.5 |
| **15-19** | 60.3 | 60.5 | 60.7 | 60.8 | 61 | 61.4 | 61.1 | 61.3 | 61.5 | 61.6 | 61.9 | 62 | 62 | 62.1 | 62.1 | 62.1 | 61.9 | 61.7 | 61.7 | 61.8 | 61.9 | 60.5 |
| **20-24** | 55.6 | 55.7 | 56 | 56.1 | 56.2 | 56.6 | 56.3 | 56.6 | 56.7 | 56.8 | 57.1 | 57.2 | 57.2 | 57.3 | 57.3 | 57.3 | 57.1 | 56.9 | 56.9 | 57 | 57.1 | 55.7 |
| **25-29** | 50.9 | 51.1 | 51.3 | 51.4 | 51.6 | 52 | 51.7 | 51.9 | 52.1 | 52.1 | 52.4 | 52.5 | 52.6 | 52.6 | 52.6 | 52.6 | 52.4 | 52.3 | 52.2 | 52.3 | 52.4 | 51.1 |
| **30-34** | 46.2 | 46.4 | 46.6 | 46.7 | 46.9 | 47.3 | 47 | 47.3 | 47.4 | 47.5 | 47.7 | 47.9 | 47.9 | 48 | 47.9 | 47.9 | 47.8 | 47.7 | 47.7 | 47.7 | 47.8 | 46.5 |
| **35-39** | 41.5 | 41.7 | 41.9 | 42 | 42.2 | 42.6 | 42.3 | 42.6 | 42.7 | 42.8 | 43.1 | 43.2 | 43.3 | 43.3 | 43.3 | 43.3 | 43.2 | 43.2 | 43.2 | 43.2 | 43.2 | 42.1 |
| **40-44** | 36.9 | 37.1 | 37.3 | 37.4 | 37.6 | 37.9 | 37.7 | 37.9 | 38.1 | 38.2 | 38.4 | 38.6 | 38.6 | 38.7 | 38.7 | 38.7 | 38.7 | 38.7 | 38.6 | 38.7 | 38.7 | 37.6 |
| **45-49** | 32.4 | 32.6 | 32.8 | 32.9 | 33.1 | 33.4 | 33.2 | 33.4 | 33.5 | 33.6 | 33.9 | 34 | 34.1 | 34.2 | 34.2 | 34.2 | 34.1 | 34.2 | 34.2 | 34.2 | 34.3 | 33.3 |
| **50-54** | 28 | 28.2 | 28.4 | 28.5 | 28.7 | 29.1 | 28.8 | 29 | 29.2 | 29.3 | 29.5 | 29.7 | 29.7 | 29.8 | 29.8 | 29.8 | 29.8 | 29.8 | 29.8 | 29.8 | 29.9 | 29 |
| **55-59** | 23.8 | 24 | 24.2 | 24.3 | 24.5 | 24.9 | 24.6 | 24.9 | 25 | 25.1 | 25.4 | 25.5 | 25.6 | 25.6 | 25.6 | 25.7 | 25.6 | 25.7 | 25.6 | 25.7 | 25.7 | 24.9 |
| **60-64** | 19.8 | 20 | 20.2 | 20.3 | 20.5 | 20.9 | 20.6 | 20.9 | 21 | 21.1 | 21.4 | 21.5 | 21.6 | 21.7 | 21.7 | 21.7 | 21.7 | 21.8 | 21.7 | 21.7 | 21.8 | 21.1 |
| **65-69** | 16.1 | 16.3 | 16.5 | 16.6 | 16.8 | 17.2 | 16.9 | 17.1 | 17.2 | 17.3 | 17.6 | 17.7 | 17.8 | 17.9 | 17.9 | 18 | 18 | 18 | 18 | 18.1 | 18.1 | 17.5 |
| **70-74** | 12.9 | 13 | 13.2 | 13.3 | 13.4 | 13.7 | 13.4 | 13.6 | 13.7 | 13.9 | 14.1 | 14.2 | 14.3 | 14.4 | 14.4 | 14.4 | 14.4 | 14.5 | 14.5 | 14.5 | 14.6 | 14.1 |
| **75-79** | 10 | 10.1 | 10.2 | 10.3 | 10.4 | 10.7 | 10.3 | 10.5 | 10.6 | 10.7 | 10.9 | 11 | 11 | 11.1 | 11.1 | 11.2 | 11.1 | 11.3 | 11.2 | 11.3 | 11.3 | 10.9 |
| **80-84** | 7.5 | 7.6 | 7.7 | 7.7 | 7.9 | 8.1 | 7.6 | 7.8 | 7.9 | 8 | 8.2 | 8.1 | 8.2 | 8.2 | 8.2 | 8.3 | 8.3 | 8.4 | 8.3 | 8.4 | 8.4 | 8.2 |

**eFigure 1.** Annual and cumulative total number of excess deaths among Black people

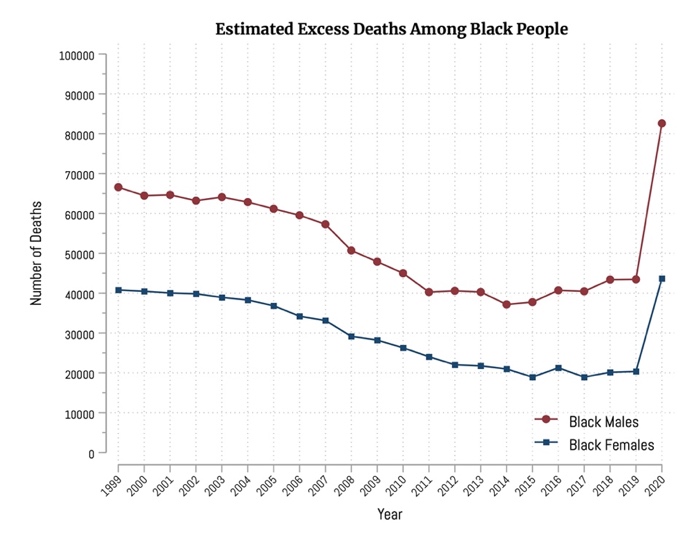

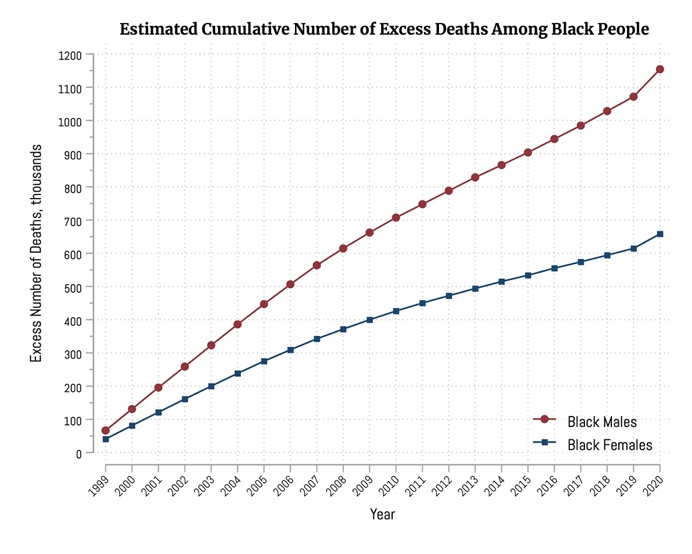

**eFigure 2**. Annual and cumulative total number of excess years of potential life lost among Black people

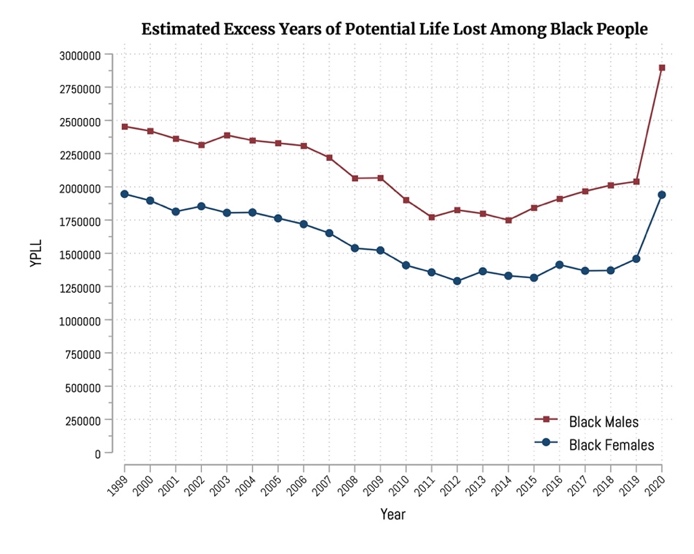

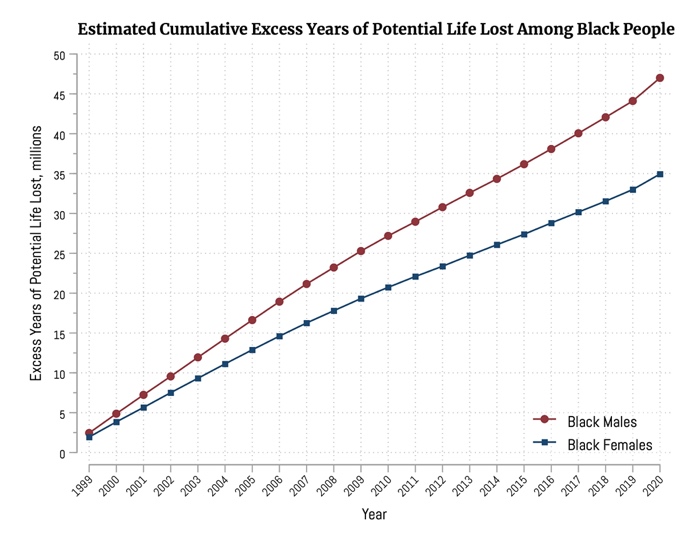
